# Enhanced Peer-Group strategies to support prevention of Mother-to-Child HIV transmission leads to increased retention in care in Uganda: A Randomized controlled trial

**DOI:** 10.1101/2023.04.15.23288495

**Authors:** Alexander Amone, Grace Gabagaya, Priscilla Wavamunno, Gordon Rukundo, Joyce Namale-Matovu, Samuel S. Malamba, Irene Lubega, Jaco Homsy, Rachel King, Clemensia Nakabiito, Zikulah Namukwaya, Mary Glenn Fowler, Philippa Musoke

**Author notes:** Corresponding author: Alexander Amone, Makerere University - Johns Hopkins University Research Collaboration, P.O. Box 23491, Kampala, Uganda. These authors have contributed equally to the work. Trial registration: ClinicalTrials.gov.

## Abstract

**Introduction:** Despite scale up of Option B+, long-term retention of women in HIV care during pregnancy and the postpartum period remains an important challenge. We compared adherence to clinic appointments and antiretroviral therapy (ART) at different follow-up time points between enrolment and 24 months postpartum among pregnant women living with HIV and initiating Option B+ randomized to a peer group support, community-based drug distribution and income-generating intervention called “Friends for Life Circles” (FLCs) versus the standard of care (SOC).

**Methods:** Between 16 May 2016 and 12 September 2017, 540 ART-naïve pregnant women living with HIV at urban and rural health facilities in Uganda were enrolled in the study. Participants were randomized 1:1 to the FLC intervention or SOC and assessed for adherence to prevention of mother to child HIV transmission (PMTCT) clinic appointments at 6 weeks, 12 and 24 months postpartum, self-reported adherence to ART at 6 weeks, 6 and 24 months postpartum validated by plasma HIV-1 RNA viral load (VL) measured at the same time points, and HIV status and HIV-free survival of infants at 18 months postpartum. We used Log-rank and Chi-Square p-values to test the equality of Kaplan-Meier survival probabilities and hazard rates (HR) for failure to retain in care for any reason by study arm.

**Results:** There was no significant difference in adherence to PMTCT clinic visits or to ART or in median viral loads between FLC and SOC arms at any follow-up time points. Retention in care through the end of study was high in both arms but significantly higher among participants randomized to FLC (86.7%) compared to SOC (79.3%), p=0.022. The adjusted HR of visit dropout was 2.5 times greater among participants randomized to SOC compared to FLC (aHR=2.498, 95% CI: 1.417 – 4.406, p=0.002). Median VL remained < 400 copies/ml in both arms at 6 weeks, 6 and 24 months postpartum.

**Conclusions:** Our findings suggest that programmatic interventions that provide group support, community based ART distribution and income-generation activities may contribute to retention in PMTCT care, HIV-free survival of children born to women living with HIV, and to the elimination of mother to child HIV transmission (MTCT).

## INTRODUCTION

Prevention of mother-to-child HIV transmission (PMTCT) remains a pillar in the global fight against HIV/AIDS. “Option B+” for PMTCT recommends lifelong triple ART for all pregnant and breastfeeding women living with HIV regardless of immune status. Option B+ also recommends protecting these women’s newborns with daily Nevirapine or Zidovudine from birth through 4-6 weeks of age, regardless of the infant feeding method (1).

In April 2012, the World Health Organisation (WHO) recommended Option B+ as the PMTCT treatment option which would provide the highest clinical benefits and programmatic advantages for both the care and treatment of mothers living with HIV and perinatal HIV prevention (1). Compared to previous proven PMTCT regimens, Option B+ was easier to deliver and more effective in reducing mother-to-child HIV transmission (MTCT) (2–4). Based on these recommendations, a number of low-income countries including Uganda adopted Option B+ as standard of care with the aim to eliminate MTCT of HIV.

Despite this progress and the proven efficacy of Option B+ (5), poor adherence to lifelong ART and low retention in care of pregnant and postpartum women living with HIV has remained an important programmatic challenge to the implementation of Option B+ especially in low resource settings (6–11). A review of studies done between 2000-2020 identified barriers to taking ART among pregnant women that included financial constraints limiting access to food and transport, and side effects of the therapy (12). Additionally, high dropout rates from PMTCT care have been documented after delivery and at two years of follow-up among mothers living with HIV (13–15), suggesting that service integration and linking mothers to routine ART services were important determinants of retention in care. These findings highlighted the critical need for innovative interventions to promote retention in HIV care and adherence to ART for the successful implementation of option B^+^.

In Uganda, adherence counseling by clinic-based midwives trained in counseling and the use of “buddies” as well as family involvement are recommended by the Ministry of Health (MOH) to support maternal ART adherence and retention in care (16). However, there is limited focus on the use of community-based support interventions. Studies in other resource-limited settings have shown better PMTCT intervention adherence with the use of community-based counseling, support groups and home visits (17–20). Additionally, studies in Kenya have demonstrated that peer support from community-based mentor mothers to improve ART adherence and retention in care was well accepted among women living with HIV (21, 22).

Given these challenges, we conducted a randomized controlled trial (RCT) to compare adherence to PMTCT clinic appointments and ART at different time points within 24 months of postpartum follow-up among women started on ART while pregnant and randomized to an enhanced peer group support intervention including community-based drug distribution and income-generating activities called “Friends for Life Circles” (FLCs) versus the MOH standard of care (SOC).

## METHODS

We conducted an open-label RCT between 16 May 2016 and May 2020 at Mulago National Referral Hospital and two health centers (HCs III & IV) in Kampala, the capital city of Uganda; and at rural Mityana District Hospital (74 kms northwest of Kampala) as well as five HCs III and IV in Mityana district. Participants were recruited from 16 May 2016 to 12 September 2017 and follow up was completed on 26 May 2020. The selected HCs represent different levels of maternal health care offered in Uganda. The study was implemented and coordinated by the Makerere University–Johns Hopkins University (MUJHU) Research Collaboration based at Mulago Hospital in Kampala.

### Enrollment and randomization procedures

We enrolled pregnant women living with HIV initiated on PMTCT Option B+ who were: ≥ 18 years old, confirmed pregnant by urine dipstick, confirmed HIV-positive by rapid HIV testing as per National PMTCT Guidelines, being ART-naïve and beginning Option B+ ART within 30 days of enrolment into the study, residing within 20 km of the study clinic, not planning to move out of the clinic catchment area within 2 years, agreeing to be home visited, and providing written informed consent.

Pregnant women testing HIV-positive through the routine National PMTCT Program and starting Option B+ were referred to study counselors by the clinic PMTCT counselors for prescreening and scheduled to return to the study clinics for consent procedures within one month of starting once daily Option B+ ART. HIV rapid testing followed the national and WHO algorithms (23) using three sequential antibody rapid tests: Abbott Determine Rapid HIV 1 Test, Abbott Laboratories, USA; Stat Pak Rapid HIV-1 Test, Chembio Diagnostic Systems, USA; and Unigold HIV Rapid Test, Trinity Biotech, Ireland.

Eligible and consenting women were then randomized 1:1 irrespective of the study sites to SOC control arm or FLC intervention arm. The randomization list was computer-generated by the data manager based at MUJHU using random-sized block groups that included consecutive intervention numbers with corresponding random intervention assignments. Each pregnant woman was assigned unique participant identification number and authors had no access to information that could identify individual participants during or after data collection

After randomization, participants were assessed for socioeconomic status, ARV drug adherence and stigma. A questionnaire addressing participants’ experiences with stigmatization and discrimination at family and community levels was administered at baseline, 12 months post-enrolment and at end of study. An additional needs assessment questionnaire was administered to participants enrolled in the FLC arm at enrolment, 1-year post enrolment and every 6 months thereafter until the end of study. This was to assess individuals’ achievements and to document challenges related to group activities and participation in income-generating activities (IGAs) and its benefits, skills acquired and training needs.

### FLC Intervention and SOC controls

The FLC intervention (hereafter ‘FLC’) included community-based enhanced peer group support, IGAs and community-based drug distribution. Participants in the FLC arm formed dynamic peer support groups based on women’s home addresses documented by Geographical Positioning System (GPS). Groups consisted of 8-10 participants who attended monthly meetings in their communities. Sustainable livelihood study assistants and study counselors participated in and documented FLC group activities. Each FLC group was supported by one peer mother who had similar social background and/or life experiences as FLC participants.

During monthly FLC group meetings, study counselors distributed ART and Cotrimoxazole and offered psychosocial support on follow-up clinic visits and drug adherence through individual and group counseling while emphasizing peer support among participants.

FLC groups also offered skills building in a variety of IGAs such as goat rearing, bakery, crafts making, liquid soap making, book binding and charcoal briquette making. These were chosen by the participants and implemented as a group or as individuals with the guidance of a study IGA Advisor. All FLC groups were linked to local government bodies for sustainability, registration and access to Community Development Driven grants funded by the District Local Government to support the implementation of livelihood projects for community-based groups.

SOC arm participants received counseling from clinic PMTCT counselors and collected their drug refills on an individual basis from the PMTCT clinics where they were enrolled as per MOH guidelines (16). SOC arm participants participated voluntarily in family support groups whenever available at their respective health facility as recommended by the MOH (16).

### Follow-up procedures

At each study site, all study participants were followed at a dedicated clinic for all scheduled and unscheduled visits and were given transport reimbursement for scheduled study visits. Scheduled visits included a first visit within the initial four weeks post-enrolment, then monthly thereafter until delivery, at delivery, at 6 and 14 weeks postpartum, and quarterly thereafter from 6 months through 24 months postpartum (end of follow-up). At each of these visits, FLC participants were counseled by study counselors on drug adherence and family planning while SOC participants were counseled by PMTCT counselors. Study counselors administered questionnaires on ART adherence, stigma, and socio-economic status to all participants at each scheduled clinic visit.

Participants were terminated from the study if they missed four or more consecutive scheduled study visits, withdrew consent, relocated outside study area, or died.

### Infant procedures

All infants born to study participants were enrolled in the study and received daily Nevirapine syrup from birth up to 6 weeks of age regardless of the infant feeding method. Infants were tested routinely at 6 weeks of age for HIV by DNA PCR and at 18 months of age by rapid HIV antibody test as part of the National Early Infant HIV Diagnosis Program (23).

### Outcomes and measures

The primary outcomes of interest included adherence to PMTCT clinic appointments at 6 weeks, 12 and 24 months postpartum and adherence to Option B+ ART at 6 weeks, and 6 and 24 months postpartum.

Complete adherence to visits was defined as having attended the 4 scheduled quarterly visits at the end of each of the first and second years of postpartum follow-up as defined by the MOH (16). Scheduled visits were determined to have been adhered to on the basis of a completed visit within a 3-week window on either side of the scheduled visit date.

Retention in care was defined as the proportion of participants who were not terminated from the study prior to the end of follow-up (24 months postpartum). Causes for termination included participant relocation out of the study area, withdrawal from the study, loss to follow-up, miscarriage, referral outside the study facility, and death. The study defined loss to follow-up as not attending 3 consecutive scheduled study visits. Retention in care was calculated as 1 minus the proportion of women terminated for any such reason before the end of follow-up.

As a second primary outcome, we compared maternal ART adherence using self-report at 6 weeks and 6 and 24 months postpartum among all study participants. We validated self-reports by comparing VL measurements for all participants at 6 weeks, 6 and 24 months postpartum.

All participants were asked to self-assess the number of complete ART doses they missed in the past three days. Self-reported ART adherence was then calculated as 1 minus the proportion of self-reported doses missed in the past 3 days.

Laboratory measures of ARV adherence were used to validate self-reports using viral suppression below 400 copies/mL. At the time of writing the study protocol, this limit was considered as viral suppression by Uganda MOH (16). VL measurements were performed at the Infectious Diseases Institute (IDI) Core Laboratory, Kampala, Uganda, a central laboratory certified by the American College of Pathologists. Nucleic acids were extracted and tested for HIV-1 RNA using the COBAS® AmpliPrep/ COBAS® TaqMan® HIV-1 Test procedure following the manufacturer’s instructions (Roche Molecular Systems, Inc., Branchburg, NJ, 08876 USA) (25).

Virological failure was defined as median viral load persistently above 1000 copies/ml for two consecutive viral load measurements within a three-month interval after at least 6 months of initiating ART. The final drug adherence variable at each time point was dichotomized as ‘not adhering’ if viral load was ≥400 copies/ml irrespective of the level of self-reported adherence, or as ‘adhering’ if self-reported adherence in the last 3 days was <u>≥</u>95% and viral load was <400 copies /ml.

Lastly, the HIV status and the HIV-free survival of children born to study participants were assessed at their 6 week and 18-month postpartum visits as per the National PMTCT program recommendation and practice.

### Statistical Analyses

We estimated a target sample size of at least 270 women living with HIV needed to be enrolled in each of the study arms to provide 90% power at 5% significance level to detect a difference of 15% or more in adherence to visits and drugs or in virological suppression between the intervention and control arms. Follow-up time was calculated in person-years, with each participant contributing time in years from enrolment to censoring at the date when a participant was lost to follow-up, died or reached the end of study.

We compared participants’ baseline characteristics in the FLC and SOC arms by testing for differences in proportions using Fisher’s exact p-values. Maternal adherence to at least 4 scheduled PMTCT clinic appointments at the end of each year of postpartum follow-up, adherence to ART and retention in care, as well as the HIV status of children were also summarized using proportions and chi-square p-values. We used the Wilcoxon–Breslow–Gehan test to assess the difference in HIV-free survival of participants’ children between the FLC and SOC arms. Log rank and Chi-Square p-values were used to test the equality of Kaplan-Meier survival probabilities and hazard rates for failure to retain in care for any reason by study arm. Cox Proportional Hazard regression model adjusted for unbalanced baseline variables was used to estimate the hazard rate ratio of failing to retain participants in care with 95% confidence intervals between the FLC and the SOC arms.

### Ethical and regulatory considerations

The study was approved by Uganda’s National Council of Science and Technology (UNCST), the Joint Clinical Research Centre (JCRC) Institutional Review Board (IRB), as well as the IRBs of Johns Hopkins University (JHU) and the University of California San Francisco (UCSF). Written informed consent was obtained from all study participants by study counselors in the presence of an impartial witness before enrolment in the study.

## RESULTS

Between May 2016 and September 2017, a total 1192 HIV-positive pregnant women were assessed for study eligibility. Of these, 45 % (540/1192) were found eligible and randomized 1:1 to either the intervention or the control arm until each arm accrued 270 participants across all study sites (Fig 1).

**Figure 1.**
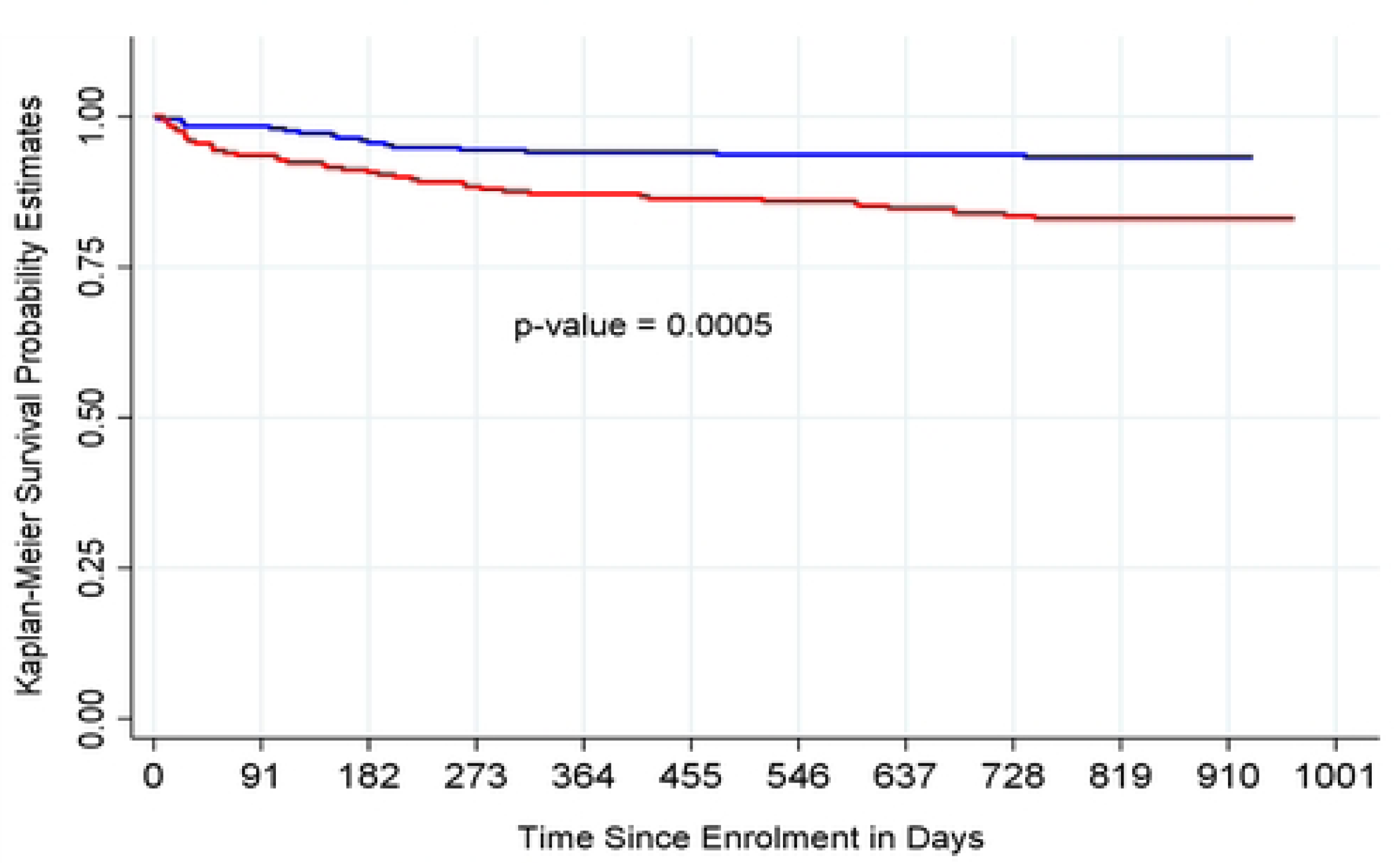
– Trial Profile.

### Baseline demographic characteristics

Table 1 provides the number of participants, clinic visits made, and person-years of follow-up by baseline socio-demographic characteristics of the study population stratified by study arm. These results show that despite randomization, there were significant differences between the two study arms in participants’ educational level, gravidity, HIV status disclosure to partner and monthly income.

**Table 1.**
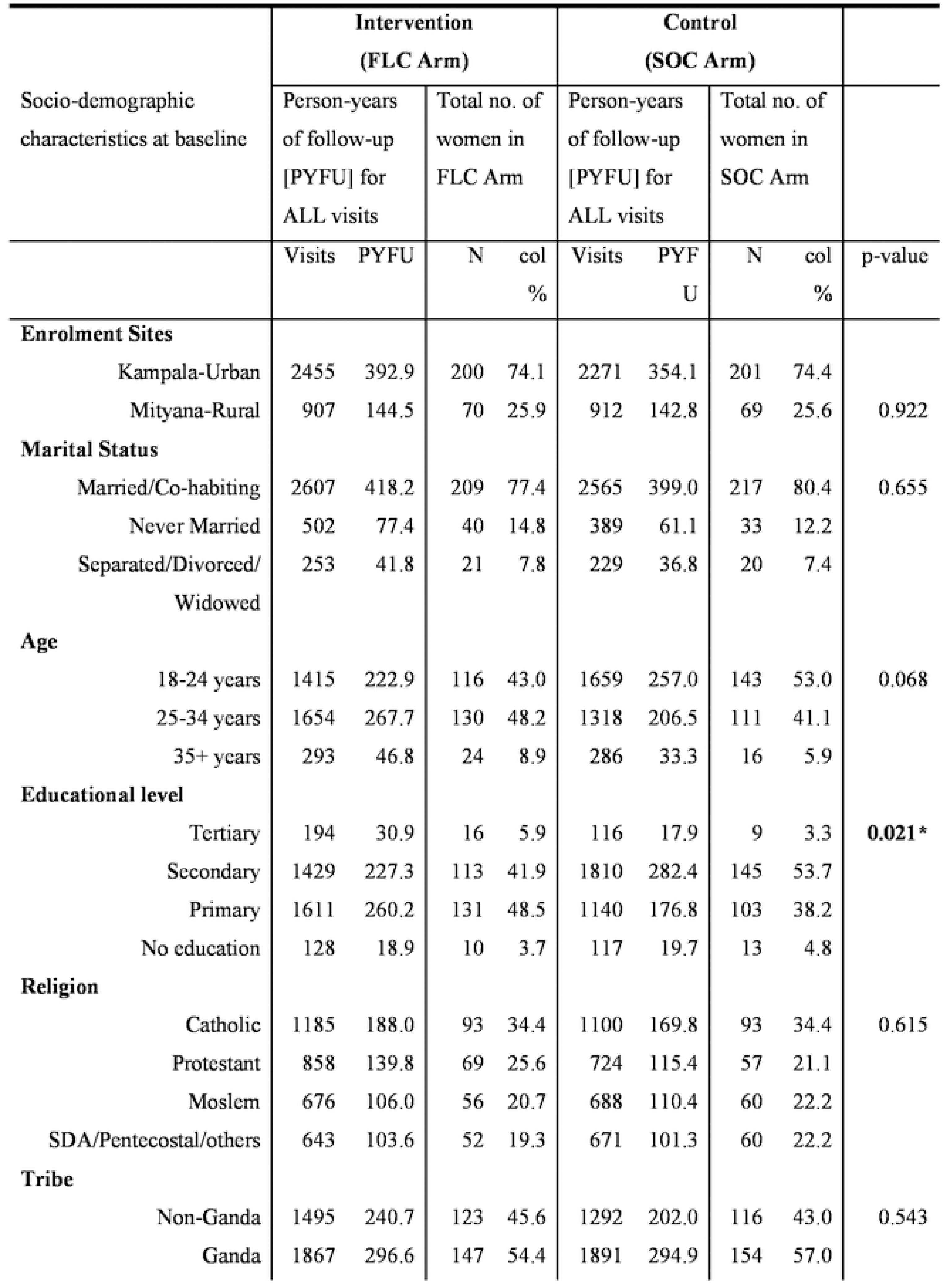
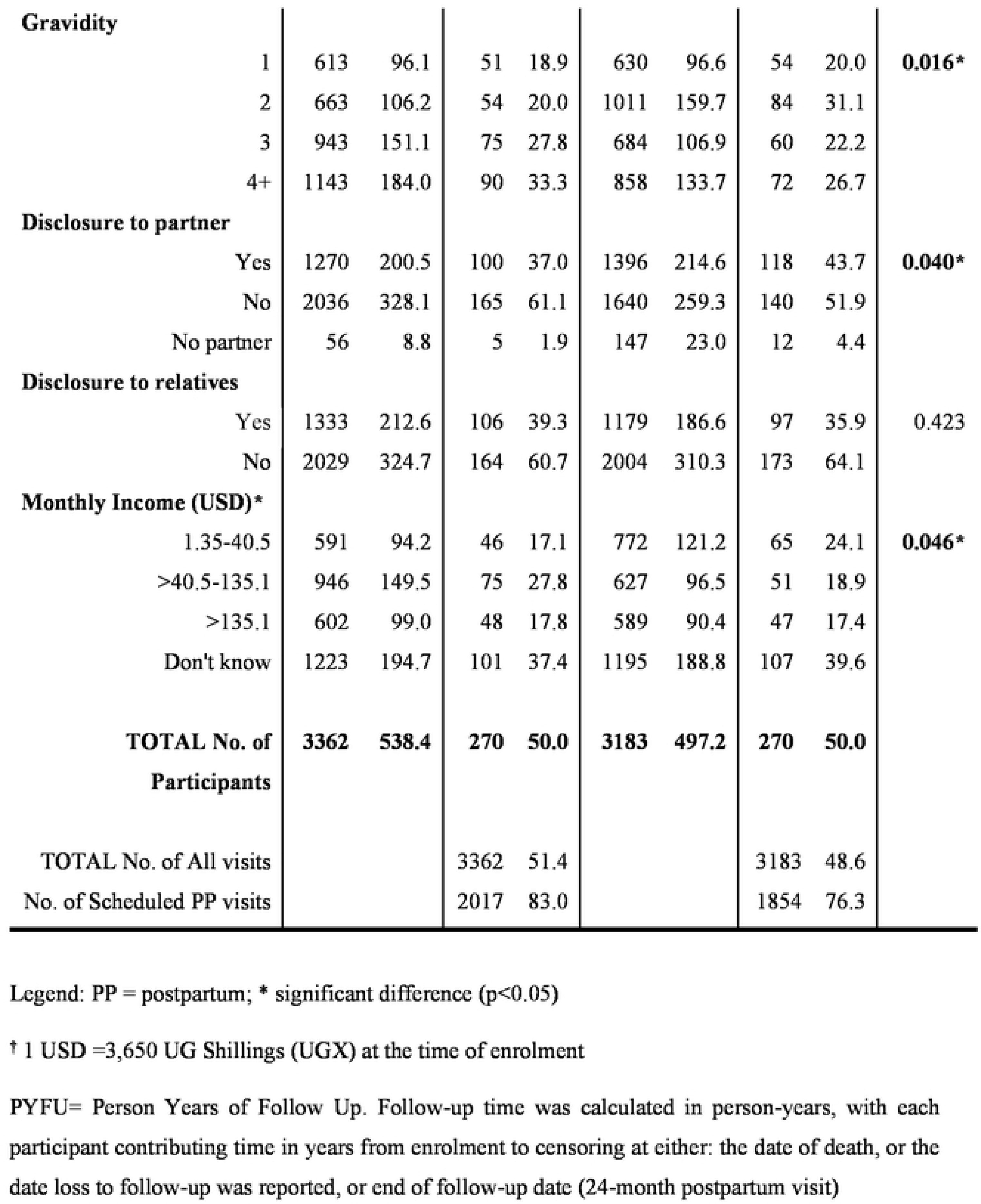
Baseline socio-demographic characteristics of study population (N=540)

### Adherence to PMTCT clinic appointments

There was no statistically significant difference between the intervention and control arms in the number of women who started or completed their postpartum visits at 6 weeks postpartum or in the first or second year of follow-up (Table 2).

**Table 2.** Adherence to PMTCT clinic appointments.

| Adherence to PMTCT clinic appointments<br>*Primary outcomes measure (A)* | At 6 Weeks Postpartum visit |  | Number attended at least 4 quarterly visits in the 1<sup>st</sup> year postpartum measured at the 12 Months Postpartum visit |  | Number attended at least 4 quarterly visits in the 2<sup>nd</sup> year postpartum measured at the 24 Months Postpartum visit |  |
| --- | --- | --- | --- | --- | --- | --- |
|  | FLC ARM | SOC ARM | FLC ARM | SOC ARM | FLC ARM | SOC ARM |
| <u>Intention To Treat analysis:</u><br>Includes all women who started the time window of interest (6 weeks postpartum, the first or the second year of postpartum follow-up) | 242/270<br>(89.6%) | 231/270<br>(85.6%) | 223/262<br>(85.1%) | 197/247<br>(79.8%) | 187/245<br>(76.3%) | 168/224<br>(75.0%) |
| Fisher's Exact p-values | P=0.192 |  | P=0.129 |  | P=0.748 |  |
| <u>As treated analysis:</u><br>Includes all women who completed the time window of interest (6 weeks postpartum, the first or the second year of postpartum follow-up) | 242/262<br>(92.4%) | 231/247<br>(93.5%) | 223/245<br>(91.0%) | 197/224<br>(87.9%) | 187/229<br>(81.7%) | 168/207<br>(81.2%) |
| Fisher's Exact p-values | P=0.730 |  | P=0.294 |  | P=0.903 |  |

PP = postpartum
<sup>a</sup> Women who attended $\geq 4$ quarterly visits in the 1<sup>st</sup> year PP
<sup>b</sup> Women who attended $> 4$ quarterly visits in the 2<sup>nd</sup> year PP

### Retention in care

Overall, retention in care was high across both study arms with 83.0% of all participants remaining in care at the end of follow-up (Table 3). Significantly, more women were retained in care at the end of follow-up in the FLC arm (86.7%) compared to the SOC arm (79.3%, p= 0.0221). Also, more women were terminated before the end of follow-up due to relocation in the SOC arm (n=27, 10.0%) compared to the FLC arm (n=12, 4.4%).

**Table 3.**
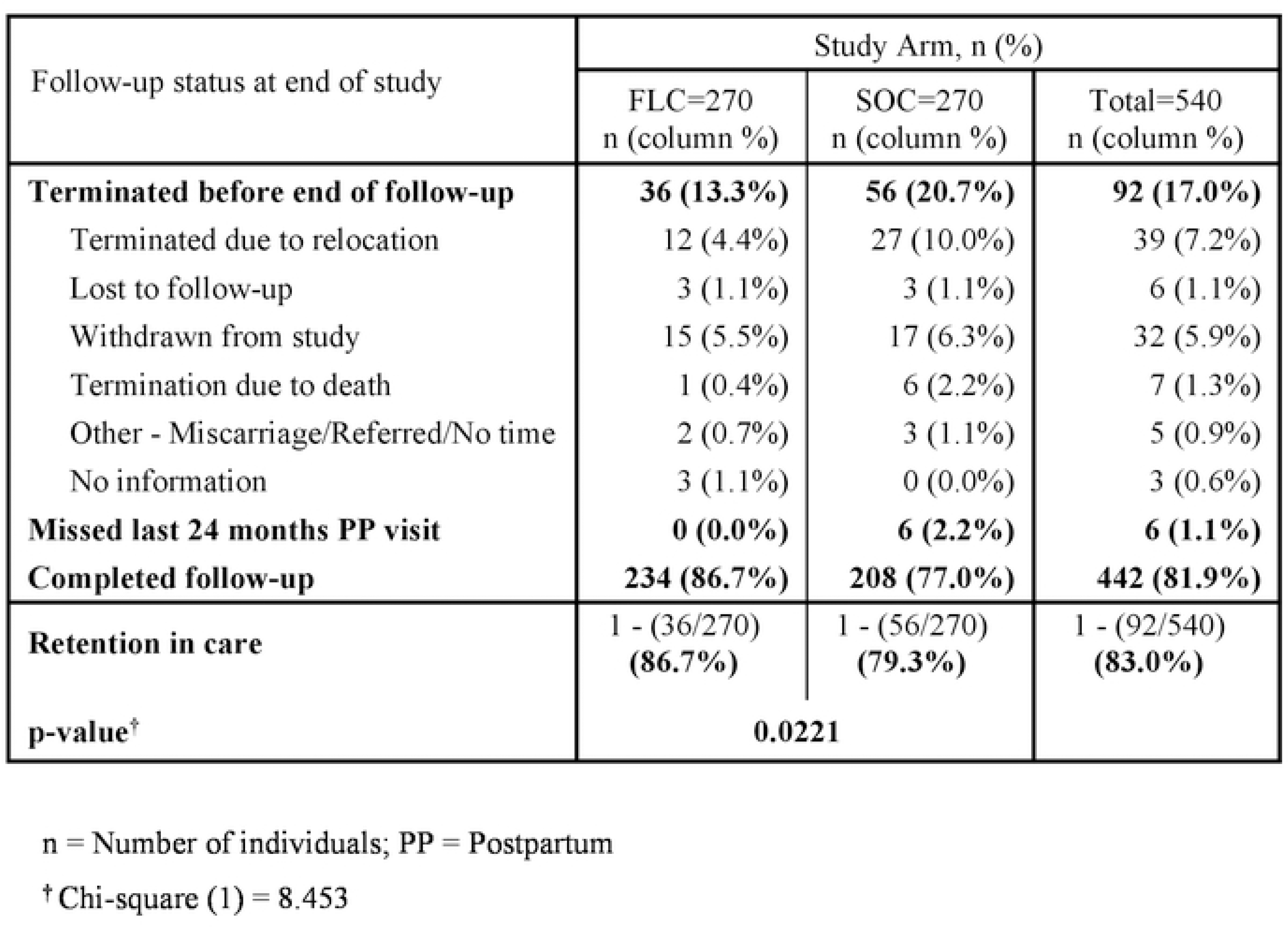
Retention in care by study arm at the end of study.

There was a significant difference between the Kaplan-Meier survival functions for failure to remain in care until the end of study (24 months postpartum visit) between the control and intervention arms (Wilcoxon-Breslow test for equality of survivor functions Chi^2^ (1) = 11.94, p-value = 0.0005) (Figure 2). Overall, the estimated proportion of participants retained in care was high, with 87.0% of all participants remaining in care until the end of the study.

**Figure 2.**
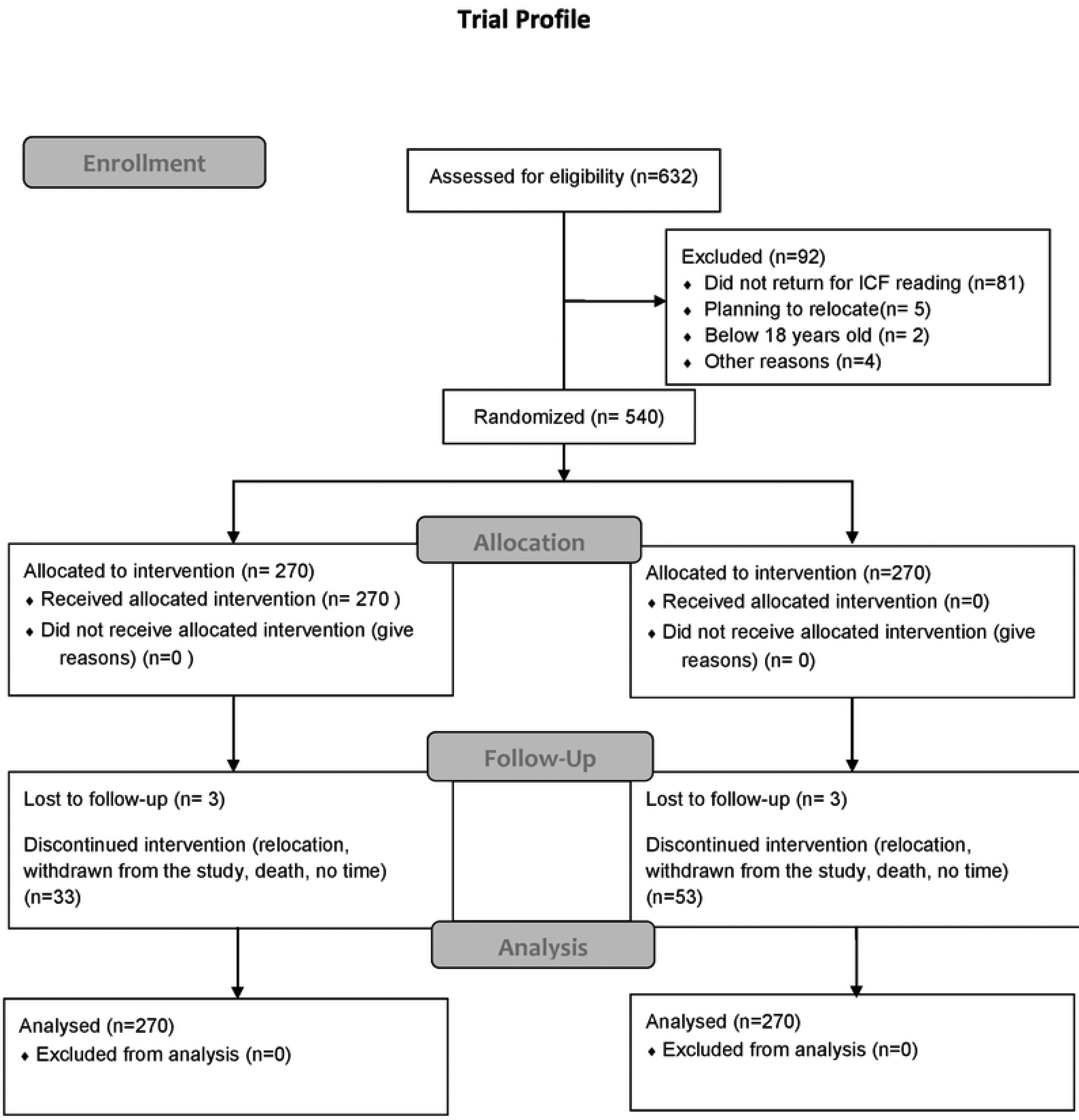
Kaplan-Meier survival curves for retention in care up to 24 months postpartum.

### Adherence to ART

Table 4 shows that participants self-reported optimal adherence to taking >95% of their ART medication in the last 3 days at their 6 weeks, 6 and 24 months postpartum visits. There was no statistically significant difference in optimal adherence to ART between the arms. The median viral loads of participants at each of these time points were <100 copies/ml with no statistically significant differences between the two arms.

**Table 4.**
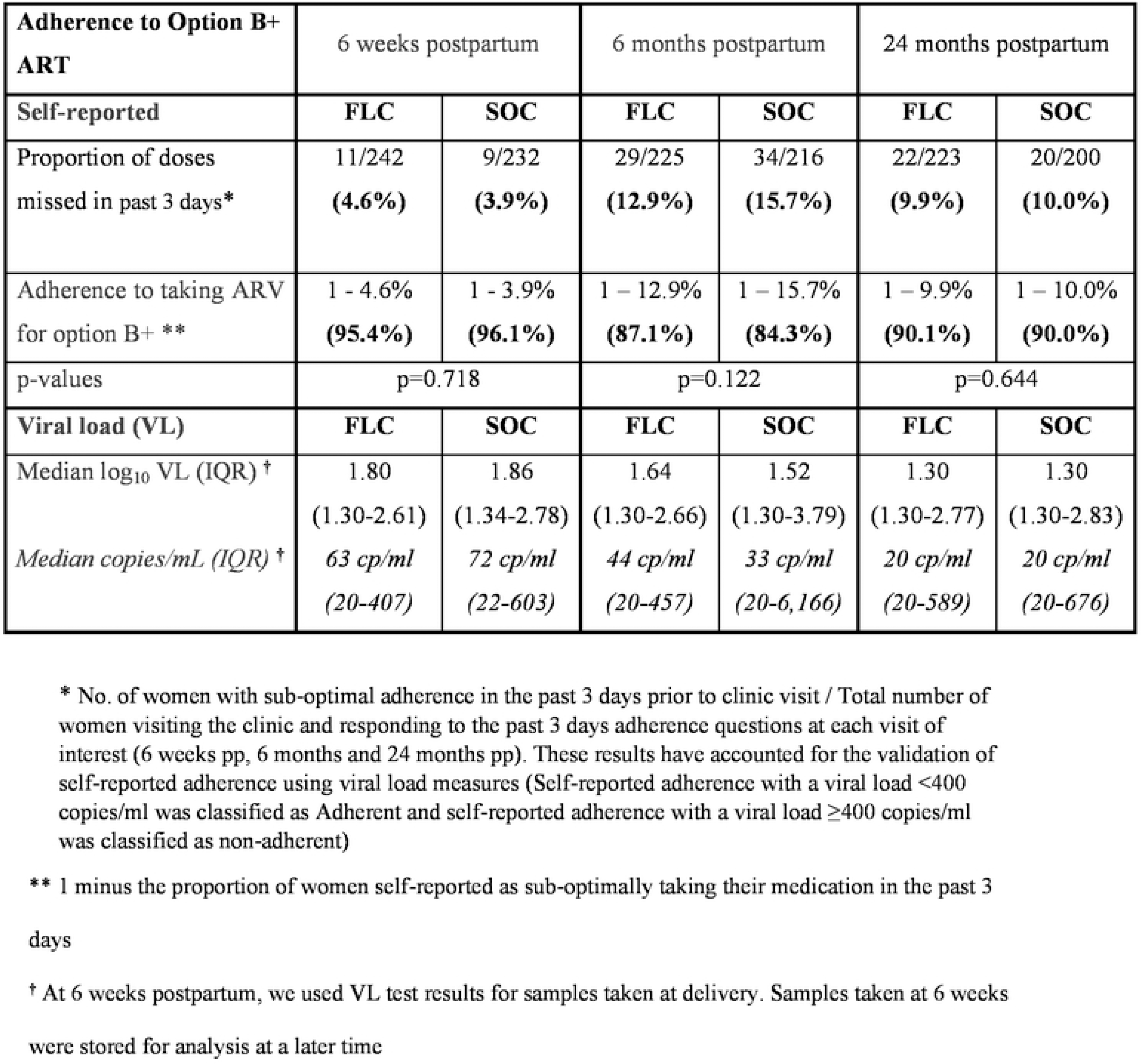
Adherence to Option B+ ART by study arm.

| Adherence to Option B+ ART | 6 weeks postpartum |  | 6 months postpartum |  | 24 months postpartum |  |
| --- | --- | --- | --- | --- | --- | --- |
| Self-reported | FLC | SOC | FLC | SOC | FLC | SOC |
| Proportion of doses missed in past 3 days* | 11/242<br>(4.6%) | 9/232<br>(3.9%) | 29/225<br>(12.9%) | 34/216<br>(15.7%) | 22/223<br>(9.9%) | 20/200<br>(10.0%) |
| Adherence to taking ARV for option B+ ** | 1 - 4.6%<br>(95.4%) | 1 - 3.9%<br>(96.1%) | 1 – 12.9%<br>(87.1%) | 1 – 15.7%<br>(84.3%) | 1 – 9.9%<br>(90.1%) | 1 – 10.0%<br>(90.0%) |
| p-values | p=0.718 |  | p=0.122 |  | p=0.644 |  |
| Viral load (VL) | FLC | SOC | FLC | SOC | FLC | SOC |
| Median log<sub>10</sub> VL (IQR) † | 1.80<br>(1.30-2.61) | 1.86<br>(1.34-2.78) | 1.64<br>(1.30-2.66) | 1.52<br>(1.30-3.79) | 1.30<br>(1.30-2.77) | 1.30<br>(1.30-2.83) |
| Median copies/mL (IQR) † | 63 cp/ml<br>(20-407) | 72 cp/ml<br>(22-603) | 44 cp/ml<br>(20-457) | 33 cp/ml<br>(20-6,166) | 20 cp/ml<br>(20-589) | 20 cp/ml<br>(20-676) |

\* No. of women with sub-optimal adherence in the past 3 days prior to clinic visit / Total number of women visiting the clinic and responding to the past 3 days adherence questions at each visit of interest (6 weeks pp, 6 months and 24 months pp). These results have accounted for the validation of self-reported adherence using viral load measures (Self-reported adherence with a viral load <400 copies/ml was classified as Adherent and self-reported adherence with a viral load ≥400 copies/ml was classified as non-adherent)
\*\* 1 minus the proportion of women self-reported as sub-optimally taking their medication in the past 3 days
† At 6 weeks postpartum, we used VL test results for samples taken at delivery. Samples taken at 6 weeks were stored for analysis at a later time

### Predictors of retention in care

Cox-proportional hazard regression models were used to assess the effect measure of intervention on being retained in care. All baseline variables with p<0.1 at the univariate level were included in the multivariate model and using a backward elimination method, only those variables with p<0.05 were kept in the final model.

Table 5 shows that the rate of dropout from care was 2.5 times greater among participants randomized to the SOC arm versus the FLC arm (aHR=2.498, 95% CI: 1.417 – 4.406, p=0.002). Participants enrolled from rural health facilities and those aged 15-24 years were also significantly less likely to be retained in care compared to their urban (p=0.021) or their older counterparts (p=0.035) respectively. Educational level, gravidity, disclosure status and income were not significant predictors of retention in care and were not included for analysis in the final model.

**Table 5:**
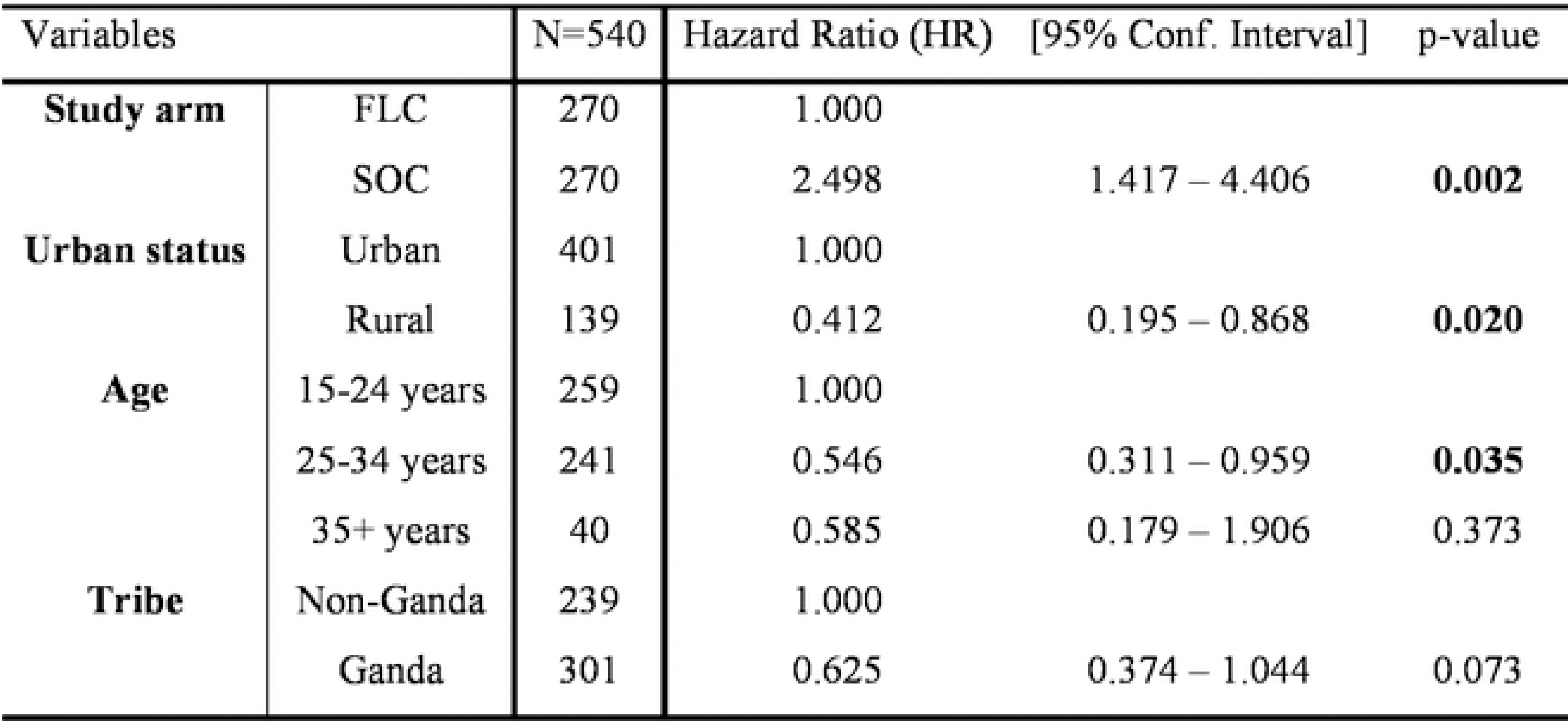
Multivariate Cox proportional hazard model for failure to remain in care.

### HIV-free survival of children

Of 492 women with a visit at 18 months postpartum, 8 of their 431 (1.9%) tested infants tested positive for HIV. Most (7/8) of these infants had tested positive to HIV by the 6th week postpartum visit. The proportion of HIV-positive children born to mothers enrolled in the FLC arm (1.4%) was not significantly different from that observed in the SOC arm (2.4%) (p=0.472).

By the 18-month postpartum visit, the HIV-free survival of children born to mothers in the FLC arm was 93.8% [95% CI: 89.9 – 96.2] compared to 83.0% [95% CI: 75.9 – 87.7] among children born to mothers in the SOC arm. The difference between the two arms was significant (p=0.0031).

## Discussion

In this RCT, we found that there was no statistical difference through 24 months of postpartum follow-up in adherence to PMTCT clinic visits or ART among pregnant and postpartum women started on Option B+ ART during pregnancy and randomized to the FLC intervention or the MOH SOC services. We however found that women in the FLC arm were statistically significantly more likely to have remained in PMTCT care through 2 years postpartum compared to the SOC arm.

This is comparable to the study by Masereka et al, which found that 87.9% of women were retained in care after being initiated on Option B+ in Uganda (26). Our data is also consistent with a study from Malawi which found that at 24 months after initiation of Option B+ ART, retention in the peer group models was 80% and 83% in the facility-based and community-based arms respectively, compared with 60% in the standard of care arm (17). This may explain the absence of a statistically significant difference between the intervention and control arm when it came to adherence to PMTCT clinic appointments.

In both arms of this RCT, 87% or more participants self-reported optimal adherence to taking their ART at 6 weeks, 6- and 24-months postpartum and the median viral load levels of participants were under 100 copies/ml at each of these time points with no significant difference between the FLC and SOC arms. We did not find fluctuations in longitudinal adherence during the postpartum period.

A systematic review by Wubneh et al in Ethiopia found that the pooled estimate of good adherence to Option B+ ART among 1852 pregnant and lactating women was 84% (27), which is comparable to our finding. Our finding is also consistent with a cohort study done in Malawi that found 90%-100% adherence to ART among women on Option B+ observed from 4 to 21 months postpartum (28). That study used pharmacy records to measure adherence as opposed to self-reports and VL validation which may limit comparisons with our study.

Although we found high self-reported adherence to ART that corroborated with low median viral loads, we did not find significant differences in these outcomes between the FLC and SOC arms at any follow-up time points. The close correlation between these outcomes echo the findings of a study among 452 women on PMTCT care and treatment in South Africa where it was found that a raised viral load was consistently associated with lower median adherence scores (29). However, the lack of a more distinct outcome from the FLC intervention may result from a study effect or still a ‘contamination’ between the FLC and SOC arms among both study staff and participants in the study facilities (30).

Few children were found infected with HIV postnatally and most of them were likely infected *in utero* or at birth given the timing of their positive test result. However, our finding that HIV-free survival of children born to mothers in the FLC arm was significantly higher than those in the SOC arm is noteworthy as it implies that more children died of non-HIV causes in the latter arm. Many factors may have accounted for this outcome but peer support, less variation in drug adherence and increased economic security (through IGAs) and thus the ability of mothers to better feed and care for their babies all are plausible explanations supported by an ample literature (31–37).

### Limitations

Our study’s main limitation is that our RCT was individually and not cluster randomized, which may have resulted in cross-contamination between the FLC and the SOC arms in the same clinics over the long-term follow-up period. Contamination could have also occurred when study staff attended to both FLC and SOC participants at the same clinic. Additionally, all study participants were given transport reimbursement for their study visits which may have positively influenced adherence to study visits. Lastly, the feasibility of our intervention would depend on its cost and cost-effectiveness, which we did not assess.

### Conclusions

Our study found that the FLC intervention significantly increased retention in care and HIV-free survival of children of pregnant mothers living with HIV through 24 months postpartum. These findings are encouraging and suggest that programmatic interventions such as the FLC may contribute to the goal of eliminating MTCT of HIV in Uganda. We recommend scaling up peer group support, community drug distribution and income-generation intervention strategies in the context of PMTCT.

## Authors’ contributions

JNM, IL, RK, JH, CN, ZN, MN, MGF and PM contributed to the conception and design of the study. AA and GG wrote the original draft. GR contributed to data curation. SM carried out statistical analysis. AA, GG, JH, PW, SM, RK, SM, CN and PM reviewed and edited the manuscript. All authors approved the final manuscript.

## Data Availability

All relevant data are within the manuscript and its Supporting Information files.

## Acknowledgements

We would like to thank the FLC for Option B+ study team who were vital in the collection of this data and the study participants without whom this study would not have been possible.

